# Effects of cladribine on intrathecal and peripheral B and plasma cells

**DOI:** 10.1101/2024.09.19.24313610

**Authors:** Kimberley Allen-Philbey, Sophie Stephenson, Gina Doody, Amy MacDougall, Mohammad Aboulwafaali, Francesca Ammoscato, Michael Andrews, Sharmilee Gnanapavan, Gavin Giovannoni, Sofia Grigoriadou, Alaco Hickey, David W. Holden, Helen Lock, Maria Papachatzaki, Iman Redha, David Baker, Reuben Tooze, Klaus Schmierer

## Abstract

**Background:** Cladribine is a deoxyadenosine analogue that can penetrate the blood-brain barrier. It is used to treat multiple sclerosis. However, the mechanistic understanding of the effect of this highly effective therapy on B cells and plasma cells in the central nervous system compartment is limited.

**Objectives:** The CLADRIPLAS study examined the effect of cladribine on peripheral and intrathecal B and plasma cell biology in people with MS.

**Methods:** Thirty-eight people with progressive MS ineligible for- or rejecting-treatment with licenced therapies were recruited and supplied a baseline lumbar puncture. Those exhibiting gadolinium-enhancing or new/enlarging T2 magnetic resonance imaging lesions and/or elevated neurofilament levels were offered subcutaneous cladribine (Litak®). Seven people were eligible; one person died before treatment, and only five completed the first year of treatment. Twenty-two ineligible people were willing to provide a repeat lumbar puncture twelve months later.

**Results:** The CLADRIPLAS study found no evidence of a difference in the odds of a positive cerebrospinal fluid oligoclonal band (cOCB) result between the cladribine-treated and untreated group. This is probably explained by microarray and *in vitro* studies, which demonstrated that plasmablasts and notably long-lived plasma cells are relatively resistant to the cytotoxic effect of cladribine compared to memory B cells at physiological concentrations. This was consistent with the loss of intracellular deoxycytidine kinase during antibody-secreting cell differentiation.

**Discussion:** CLADRIPLAS indicates that cOCB are not rapidly eliminated in most people with MS. This may be explained by the relative lack of direct cytotoxic action of cladribine on long-lived plasma cells.

## INTRODUCTION

Multiple sclerosis (MS) is a chronic, immune-mediated disease of the central nervous system (CNS), characterised by demyelination and degenerative changes ^1^. While the clinical course of MS is heterogeneous, accelerated brain and spinal cord atrophy of people with MS (pwMS) occurs at an early disease stage resulting in disability accrual ^2,3^. The recent success of B cell depleting therapies has led to a re-appraisal of adaptive immunity in MS, with B subsets now considered to be ‘disease-drivers’, rather than ‘passengers’ in what was previously thought to be a fundamentally T cell-mediated pathophysiological process ^4,5^. The presence of pathogenic oligoclonal immunoglobulin bands (OCB) in the cerebrospinal fluid (CSF) is a regular finding in pwMS, so much so that they contribute to the diagnostic criteria ^6,7^. OCB are thought to be produced by plasma cells, which are terminally differentiated B lineage cells ^8^.

Cladribine is an effective disease-modifying treatment (DMT) in pwMS leading to significant reduction of intrathecal neurofilament light chain (NfL) levels ^9^ and long-term depletion of the memory B cell subset ^10^. Whilst peripheral memory B cell depletion is a rather generic mechanism through which relapsing, MS-related inflammation can be controlled by DMT ^11^, cladribine is unique among the licensed memory B cell-depleting agents, which are typically CNS-excluded, as it is a small molecule penetrating the CNS at biologically active levels ^12,13^. Evidence suggests that OCB following treatment with cladribine may become undetectable, when assessed years later ^14^. It has been hypothesised that cladribine may deplete plasma cells and OCB shortly following treatment. To address this mechanistic question, we collected blood and CSF samples from pwMS enrolled into the *“Does cladribine target plasma cells and reduce neuro-axonal damage in people with MS*” (CLADRIPLAS) study. As per standard of care at our Trust, patients were screened for treatment eligibility with subcutaneous cladribine (SClad) at baseline and, if eligible, offered treatment with SClad ^15^. Eligibility was based on evidence of inflammatory activity CNS lesions on magnetic resonance imaging (MRI) and/or elevated CSF-neurofilament light chain (cNfL) levels). PwMS ineligible for SClad served as a reference group. CSF-OCB (cOCB) and cNfL, alongside blood B cell subpopulation analyses were undertaken. Moreover, we used blood samples from healthy donors to explore B cell differentiation in cell culture followed by microarray analysis and cytotoxicity assays for mechanistic studies. Our data provide new insights into how cladribine affects intrathecal OCB, and the impact of these effects on the significant and long-term efficacy of cladribine in pwMS.

## METHODS

### Ethics approvals

CLADRIPLAS was approved by the North of Scotland Research Ethics Committee 1 (Integrated Research Application System ID: 240360). The Leeds East Research Ethics Committee approved the *in vitro* studies of the effect of cladribine on the different stages of B cell differentiation (Ref: 07/Q1206/47).

### Study population and recruitment

Based on an extensive compassionate-use programme using subcutaneous cladribine (Litak®; SClad), participation in CLADRIPLAS was offered to pwMS who were considered for SClad treatment at Barts Health National Health Service (NHS) Trust. Patients enrolled were offered this off-label treatment based on the following criteria: (i) lack of eligibility for NHS approved DMT, or rejection by pwMS of the approved options offered; (ii) revision and understanding of a comprehensive information pack ^16^ aiding their understanding of the risks and potential benefits of SClad; (iii) disease-activity defined as (a) CNS MRI activity (gadolinium-enhancing (Gd^+^) lesion(s), new/enlarging T_2_ lesion(s) and/or (b) elevated cNfL levels; and (iv) treatment approval by the Barts Health MS neuroinflammation multidisciplinary team ^9,15,17^. All pwMS enrolled in CLADRIPLAS underwent at least one lumbar puncture (LP) as part of the study procedures. Those with an elevated cNfL level and/or MRI activity were offered SClad and, provided they went ahead, became part of the treatment arm. Those pwMS whose NfL level was within the reference range and with no detectable MRI activity became part of the reference arm.

### Sample size

A sample size of 20 subjects per arm (SClad-treated arm and untreated arm) was calculated. This was based on the assumption that 100% of untreated individuals would remain OCB positive after 1 year of study, while in the SClad group this may reduce to 60% (Rejdak et al. 2019). To detect a difference of this magnitude or larger with 80% power at two-sided 5% significance 16 participants per group were required. Allowing for 20% dropout, we aimed to recruit 20 in each group.

### Trial objectives and endpoints

Our primary objective was to study the impact of SClad on peripheral and intrathecal B-cell biology. The secondary objective was the effect of SClad on the cNfL level. The primary endpoint was binary, either cOCB positive or negative. The secondary endpoint was cNfL and serum NfL (sNfL).

### Clinical study procedures

Clinical assessments included the Expanded Disability Status Scale (EDSS) ^18^, Nine-hole peg test (9HPT), Symbol Digit Modalities Test (SDMT), Timed twenty-five-foot walk (T25FW) and the ABILHAND questionnaire ^19^. CSF was collected using an atraumatic LP procedure ^20^, alongside venipuncture for blood samples.

### Analysis of neurofilament light chain in cerebrospinal fluid and serum

After collection, both CSF and serum samples were preserved at −80°C. cNFL and sNfL were detected using a fully automated HD-X platform (Quanterix, Billerica, MA), using a commercially available NfL Simoa Assay Advantage kit, as per instructions from the manufacturer. The calibrators, ranging from 0-2000pg/ml, and controls were run neat. sNfL and cNfL samples were diluted 1:4/1:100 respectively. The signal from calibrators was fitted to a 1/Y2-weighted four-parameter logistic curve, which was used to calculate NfL concentrations. Intra- and interassay variabilities were <10%. The UmanDiagnostics Nf-light ELISA assay was also used to quantitatively determine cNfL.

### B cell subpopulations

The assay for peripheral blood B cell subsets (BSubs) was performed within 48 hours of peripheral blood collection using whole blood EDTA samples ^21^. Samples were read and analysed respectively using either (i) the Becton, Dickinson FACSCanto II Flow Cytometer and Diva software or (ii) the BD FACSLyric Flow Cytometer and FACSSuite software (BD Biosciences, UK). BSubs were read on the BD FACSCanto II machine until March 2021. The laboratory switched to the BD FACSLyric machine in April 2021. An acceptable level of bias for medium and high levels of BSubs and a non-significant bias for low levels of BSubs was detected, which was unlikely to have any material impact on the results reported.

### Microarray data analysis

Gene expression profiling of B cell differentiation was extracted from public databases using the www.genomicscape.com platform ^22^. This included human B cells to plasma cells GCRMA (n=38), Affymetrix Human Genome U133 Plus 2.0 microarray dataset ^23,24^ and the *in vitro* generation of long-lived plasma cells (n=21), Illumina HumanHT-12 V4.0 expression beadchip dataset ^25,26^. The distribution of *DCK* and other enzymes was assessed using mRNA expression data expressed as arbitrary units (a.u.).

### Effect of cladribine on different stages of B cell differentiation

The same *in vitro* model of B cell differentiation was used to investigate the effect of cladribine in dimethyl sulfoxide (Sigma) on B cells at different stages of their differentiation ^25,27^. Peripheral blood was obtained from 4 healthy donors. Mononuclear cells were isolated by Lymphoprep (Abbott) density gradient centrifugation. Total B-cells were isolated by a memory B cell negative selection kit (Miltenyi). Cells were maintained in Iscoves Modified Dulbecco Medium (IMDM) supplemented with Glutamax and 10% heat-inactivated foetal bovine serum (Invitrogen). On Day 0 to day 3, B cells were cultured in 24 well plates at 2.5 × 10^5^/ml with F(ab’) goat anti-human IgA, M and IgG (2µg/ml) (Jacksons ImmunoResearch) with 20U/ml interleukin two (IL-2) (Miltenyi) and 50ng/ml IL-21 (Miltenyi) on γ-irradiated CD40L expressing L-cells (6.25 × 10^4^/well). On day 3, cells were detached from the CD40L L-cell layer and reseeded at 1 × 10^5^/ml in media supplemented with IL-2 (20 U/ml) and IL-21 (50 ng/ml). Lipid Mixture 1, chemically defined and MEM amino acids solution (both at 1x final concentration) were added from day 3 onwards. On day 6 cells were harvested and seeded at 1 × 10^6^/ml in media supplemented on day 6-13, with IL-6 (10ng/ml) (Peprotech), IL-21 (10 ng/ml) and 100ng/ml multimeric APRIL/TNFSF13 (R&D).

Drug-free dimethyl sulfoxide (DMSO) was used as the baseline control. The concentrations of cladribine used were 0.0025µM, 0.05µM, 1µM and 20µM. Selected doses were based on the plasma pharmacokinetic profile of cladribine ^13^; Hermann et al. 2019). On day 3, cells were plated at a density of 1×10^5^/ml in media supplemented with IL-2 (20 U/ml), IL-21 (50 ng/ml) and the indicated doses of cladribine for 72 hours. On days 6 and 13, cells were plated at a density of 2×10^5^/ml in media supplemented with IL6 (10 ng/ml), IL-21 (10 ng/ml) (Day 6 only), APRIL (100ng/ml) and the indicated doses of cladribine for 48 hours.

Percentage cell survival was assessed by calculating the *per input B cell* value for each concentration of cladribine relative to DMSO. Similar total B cell cytotoxicity was evident following a 48-72 hour incubation.

### *In vitro* cell cytotoxicity demonstrating insensitivity of plasma cells to cladribine

Total B cells (CD19+, CD20+) were extracted from peripheral blood and expanded (day 0-3) and differentiated (day 0-6) into plasmablasts (CD27+, CD38+, CD138-) using stimulation of the B cell receptor, CD40 costimulation and cytokines (Figure 2A. 2B). The plasmablasts were then differentiated further to plasma cells (day 6-13) using APRIL, IL-6 and IL-21 and maintained as post-mitotic, long-lived plasma cells (CD38+, syndecan-1/CD138+) (Figure Figure 2A, 2C).

### Flow cytometric analysis

Cultured cells were analysed using 6-colour direct immunofluorescence staining on a Cytoflex LX (Beckman Coulter) flow cytometer. Antibodies used were: CD19-PE (LT19), CD138-APC (44F9) (Miltenyi); CD20-e450 (2H7) (eBioscience); CD27-FITC (M-T271), CD38-PECy7 (HB7). Controls were isotype-matched antibodies. Dead cells were excluded from analysis by Zombie UV fixable viability kit (Biolegend). Analysis was performed with FlowJo version 10 (BD Biosciences) and Prism 9 (GraphPad). Cells were enumerated by the addition of Countbright counting beads (Invitrogen).

### Statistical analysis

The primary endpoint was analysed by applying Fisher’s exact test comparing proportions of cOCB positive and cOCB negative samples to test for a difference between the treated and untreated groups at the second study visit. Median ratios between visits 1 and 2 in the treated and untreated groups were calculated for cNfL, lymphocyte count and memory B cells. All analyses were performed using the statistical package R v3.6.1. In addition for post-hoc analysis, differences between groups for continuous measurements were assessed using Student’s t-test or repeated measures analysis of variance using Sigmaplot V14 software.

## RESULTS

### Demographics of trial participants

A total of 38 pwMS with either primary (PPMS) or secondary (SPMS) progressive MS were recruited to the CLADRIPLAS study by January 2022 (Table 1), with the last patient visit conducted in January 2023. Twenty-seven pwMS completed the study. One/38 patients had their LP under fluoroscopic guidance. All but one participant had two study LPs; of one participant a historic CSF collected shortly before consent with a single follow-up LP during CLADRIPLAS. Ten pwMS withdrew from the study following their first study visit. The main reasons were the burden of travel (n=5) and not wishing to undergo another LP (n=5). The median time between visits for the SClad-treated group was 1.22 years (Interquartile range (IQR): 1.03, 1.25), and 1.08 for the untreated group (IQR: 1.02, 1.17). Seven pwMS were eligible for treatment with cladribine, all but one, who had new MRI brain lesions, based on an elevated cNfL. Five pwMS underwent two treatment courses of cladribine. One pwMS received one treatment course of cladribine prior to study withdrawal. One pwMS (EDSS 8) died of aspiration pneumonia after their first study visit prior to receiving a first dose of SClad. Twenty-two pwMS were not eligible for SClad due to lack of disease activity (Supplementary figure 2).

**Table 1.** CLADRIPLAS cohort demographics and baseline clinical characteristics.

| Characteristic | CLADRIPLAS Cohort, n=38 |
| --- | --- |
| Age at first study visit, years | 57.9 [47.9, 66.1] |
| Gender n, (%) | Female 19 (50.0)/ Male 19 (50.0) |
| Ethnicity n, (%) | White 33 (86.8)/ Other 5 (13.2) |
| MS course n, (%) | PPMS 14 (36.8)/SPMS 23 (60.5)/ RRMS 1 (2.6) |
| Disease duration (From first symptom), years | 19 [14.0, 29.8] |
| Received prior DMT. n, (%) | Yes 23 (60.5)/No 15 (39.5) |
| EDSS to assess disability | 6.5 [5.5, 7.0] |
| SDMT to assess psychomotor speed | 34.5 [22.5, 29.3] |
| Average 9HPT Dominant hand, seconds | 30.12 [24.9, 48.9] |
| Average 9HPT Non-dominant hand, seconds | 29.9 [24.8, 46.7] |
| T25FW, seconds | 11.63 [6.9, 22.4] |
| ABILHAND assessment | 31 [20.8, 39.3] |
| CSF-NfL Level (pg/ml) | 467.5 [344.5, 532] |

Demographics of pwMS recruited to the CLADRIPLAS study. All continuous variables are shown as median (interquartile range). CSF cerebrospinal fluid, DMT disease-modifying therapy, EDSS expanded disability status scale, 9HPT 9-hole peg test, MS multiple sclerosis. NfL neurofilament light chain. PPMS primary progressive MS, SDMT symbol digit modality test, SPMS secondary progressive MS.

### Lymphocyte depletion by cladribine

The median ratio between total lymphocyte count (TLC) between visits 1 and 2 in the untreated group was 0.98 (IQR: 0.85, 1.09). By contrast, in those treated with SClad a reduction of TLC by 35% (median ratio 0.62 (IQR: 0.47, 1.67) was detected between visits. There was a mean 48% drop in TLC compared to the untreated group (exponentiated estimate: 0.52 (95% confidence interval (CI): 0.29, 0.71). Likewise, the immunoglobulin-switched (CD19+, CD27+, IgD-) memory B cell count in the untreated group remained essentially unchanged. With a median ratio of 1.07 (IQR: 0.98, 1.17) between visits 1 and 2 they were 44% lower in the SClad-treated group (Median ratio 0.56 (IQR: 0.47, 0.64) at visit 2 compared to visit 1, indicating that treatment reduced switched-memory B cells by 48% compared to the untreated group (exponentiated estimate: 0.52, 95% CI: 0.38, 0.67). This suggests that SClad was biologically active in these PwMS.

### Analysis of primary endpoint

The intended number of pwMS eligible for treatment was not achieved (Table 2). Within the sample (n=38) there were five pwMS in the cladribine-treated group and 22 in the untreated group. It was found that 60% of the group on treatment (n=3/5) were cOCB positive at visit 2 compared to 82% of the untreated group (n=18/22). Fisher’s exact test (p=-0.30) provided no evidence that there was a significant difference, in odds of a positive result at visit 2, between the treated and untreated group indicating that cOCB are not rapidly eliminated in most pwMS.

**Table 2.** Influence of subcutaneous cladribine on CNS inflammation during CLADRIPLAS.

| Treatment | Sample | n | Visit 1 | Visit 2 | Probability |
| --- | --- | --- | --- | --- | --- |
| **CSF-Oligoclonal band positive** |  |  |  |  |  |
| Control | CSF | 22 | 22/22 | 18/22 | 0.108 |
| Cladribine | CSF | 5 | 5/5 | 3/5 | 0.444 |
| **Neurofilament Light (pg/ml)** |  |  |  |  |  |
| Control | Serum | 22 | 15.93 ± 8.41 | 15.38 ± 5.76 | 0.601 |
|  | CSF | 22 | 28.12 ± 12.81 | 25.99 ± 8.17 | 0.818 |
| Cladribine | Serum | 4 | 32.48 ± 13.08 | 30.809 ± 9.85 | 0.539 |
|  | CSF | 4 | 101.57 ± 55.19 | 59.00 ± 25.95 | 0.154 |

People with progressive MS enrolled to provide a CSF sample at baseline and 12 months later. People were allocated to cladribine with baseline evidence of disease activity based on MRI and/or cerebrospinal fluid (CSF) neurofilament light levels. The results represent the number of cOCB-positive samples and the mean ± standard deviation neurofilament light levels. Differences between visit 1 (baseline) and visit 2 (12 months), within the group, were assessed using Fisher Exact test or repeated measures analysis of variance.

### CSF-neurofilament

The median ratio between cNfL at visit 1 and visit 2 in the untreated group was 1.26 (IQR: 0.97, 1.97). That is, the median cNfL at time 2 was 26% higher compared to visit 1. The median ratio between cNfL at visit 1 and visit 2 in the treated group was 0.85 (IQR: 0.54, 1.43). That is, the median cNfL at time 2 was 15% lower compared to visit 1. In the treatment group cNfL level was reduced by, on average, 38% more compared to the untreated group (exponentiated estimate: 0.62, 95% CI: 0.31, 1.04). Whilst the CSF of neurofilaments reduced in the cladribine treated group, between visits, this failed to reach statistical significance (Table 2), consistent with study being underpowered. Any benefit could also relate to regression to the mean effects.

### Microarray data analysis

Microarray analysis of *deoxycytidine kinase* (*DCK*) gene expression from BSubs demonstrated a *DCK* reduction as B cells differentiated into plasmablasts and plasma cells (Figure 1A-E). As such the *DCK* mRNA levels were significantly higher in memory B cells (3,104 ± 1,024 a.u. Affymetrix array. Probe 203302_at. n=5) than expressed by plasmablasts (699 ± 260a..u. P<0.001. n=5) and bone-marrow plasma cells (207 ± 326. P<0.001. n=5/group). This was replicated across different datasets and microarray platforms (Figure 1F-J). This was seen in our own dataset (Figure 1F-J) using *in vitro* generation of plasma cells from total B cells.(Figure 2A). Using the Illumina array it could be seen that *DCK* mRNA levels (Probe ILMN 1651433) reduced from that expressed by memory B cells on day 0 from 955 ± 240 a.u. (n=3) to 193 ± 14 a.u. in plasma cells (Day 13) cultures (n=3) P= 0.005 ^25^ (Figure 2F). In contrast the expression of *adenosine deaminase* (*ADA*. Probe ILMN_1803686) increased from memory cells (193 ± 5.8) to Day 13 plasma cells (1,510 ± 52. P<0.001), whilst 5’ nucleotidases that could impact cladribine were comparable (*NT5C1A*. Probe ILMN_1815796. Memory cell 180 ± 11 a.u. vs. day plasma cell 180 ± 11 a.u. n=3. *NT5C1B*. Probe ILMN_1756623. Memory cell 158 ± 5 a.u. vs. plasma cell 169 ± 16 a.u. n=3 and *NT5C2*. Probe ILMN-1682165. Memory cell 1218 ± 101 a.u. v plasma cell 1347 ± 89 a.u. n=3). This suggested that B cells would become increasingly less sensitive as they differentiated from memory B cells to plasmablasts and from plasmablasts to plasma cells. Indeed this was found to be the case following *in vitro* culture.

**Figure 1.**
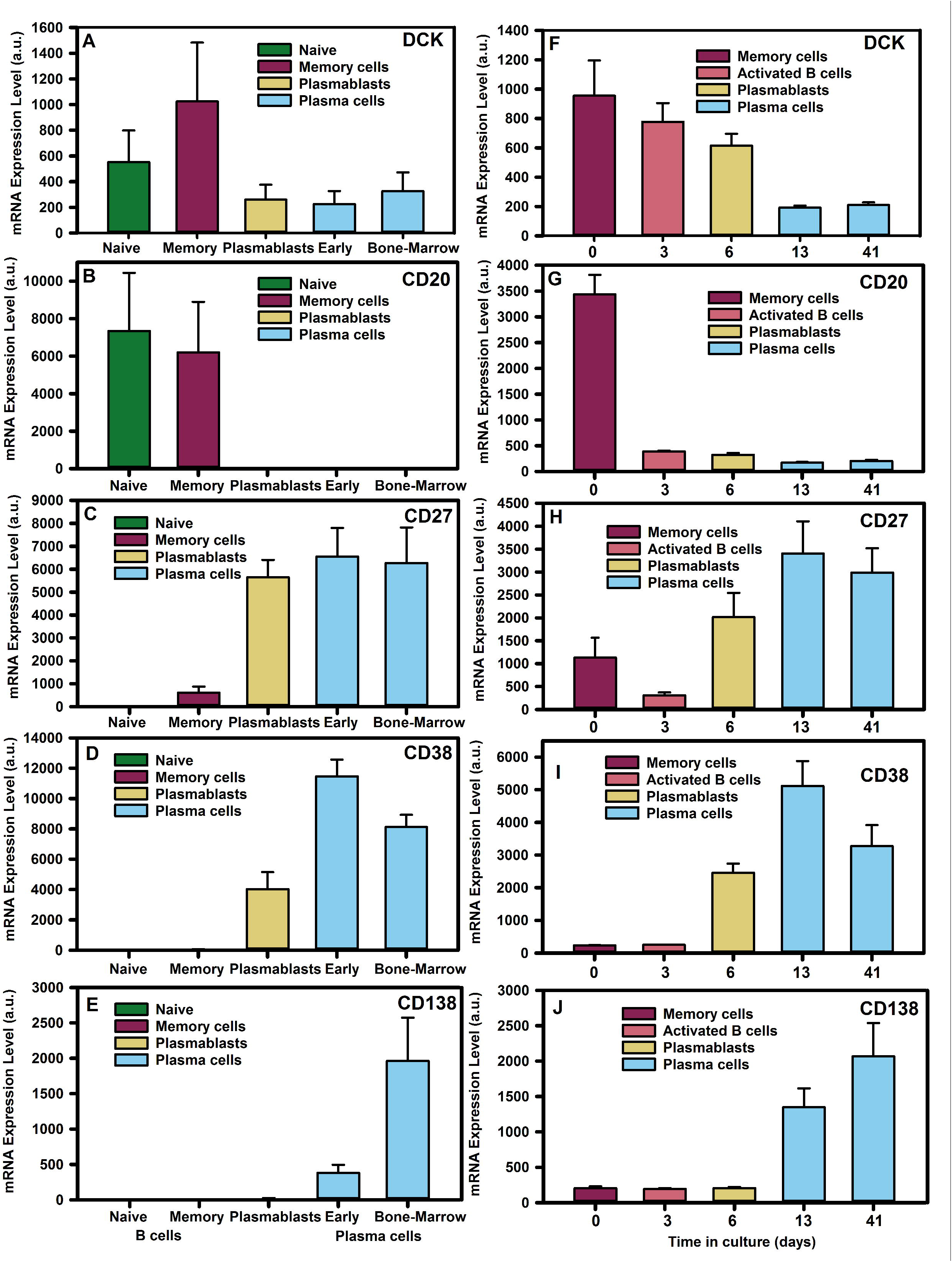
Deoxycytidine kinase is down-regulated during plasma cell development. Microarray analysis of B cell subsets from (A-E) *ex vivo* flow cytometry sorted cells, (F-J) *in vitro* differentiated plasma cells derived from total B cells using (A-E) Affymetrix human genome U133 plus 2.0 Array or the (F-J) Illumina Human-HT V4.0 expression bead chip detecting: deoxycytidine kinase (DCK. Probes 203302_at and ILMN_1651433), membrane spanning four A one (CD20. Probes 210356_x. ILMN_1776939), TNFRSF7 (CD27 probes 206150_at, ILMN _1688959), (CD38. Probes 205692_at, ILMN_2233783), Syndecan-one (CD138. Probes 20186_at, ILMN_1815303). Data was extracted from www.genomicscape.com database and the results represent the mean + standard deviation mRNA expression (arbitrary units (a.u.)) from (A-E) n=5/group or (F-J) n=3/group.

**Figure 2.**
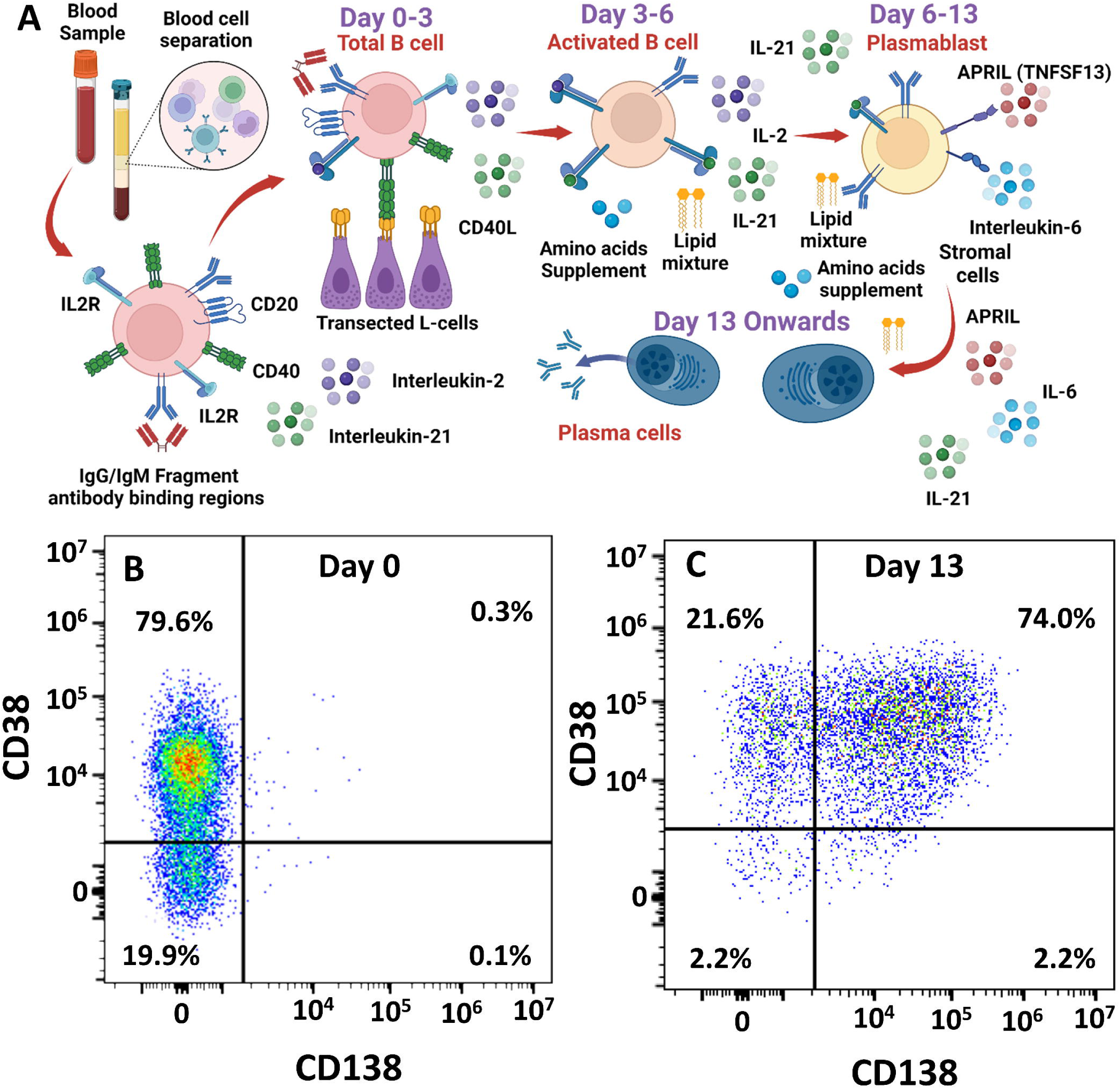
*In vitro* differentiation of B cells into plasma cells. **(A)** Peripheral blood mononuclear cells were collected and **t**otal circulating B **c**ells were purified. These were incubated with supplements, cytokines and stimulatory and co-stimulatory signalling elements to generate long-lived plasma cells. Created with Biorender.com (**B-C**) Flow cytometry demonstrating plasma cell differentiation of B cells from: (**B**) total B cells on day 0 to (**C**) plasma cells on day 13 in culture using staining with antibodies against CD38 and CD138. The results show the fluorescence intensities in representative samples.

Cells were incubated with various concentrations of cladribine during the total B cell to plasma cell transition. The doses of SClad used in CLADRIPLAS were selected to be equivalent to oral cladribine ^9^, which exhibits a 95 percentile Cmax at approximately 0.4µM and about 25% brain penetration ^13,28,29^. The *in vitro* doses examined therefore encompassed the range of *in vivo* concentrations of drug encountered. The B cells were differentiated (Figure 2A) and cell viability was assessed following incubation of the cells with various concentrations of cladribine (Figure 3). It was evident that differentiating memory B cells were highly susceptible to killing by 0.05µM cladribine, which was near complete a 1µM (Figure 3), as seen previously (Ceronie et al. 2018). This led to the elimination of cells within the lymphocyte gates during flow cytometry (Supplementary figure 1). There was however limited killing of differentiating plasmablasts at 0.05µM but again marked cytotoxicity at 1µM. In contrast there was limited cytotoxicity of plasma cells at 1µM although 20µM induced some cytotoxicity (Figure 3). This supports the view that plasma cells are relatively resistant to cladribine mechanistically supporting the cladribine-related biology observed *in vivo*.

**Figure 3.**
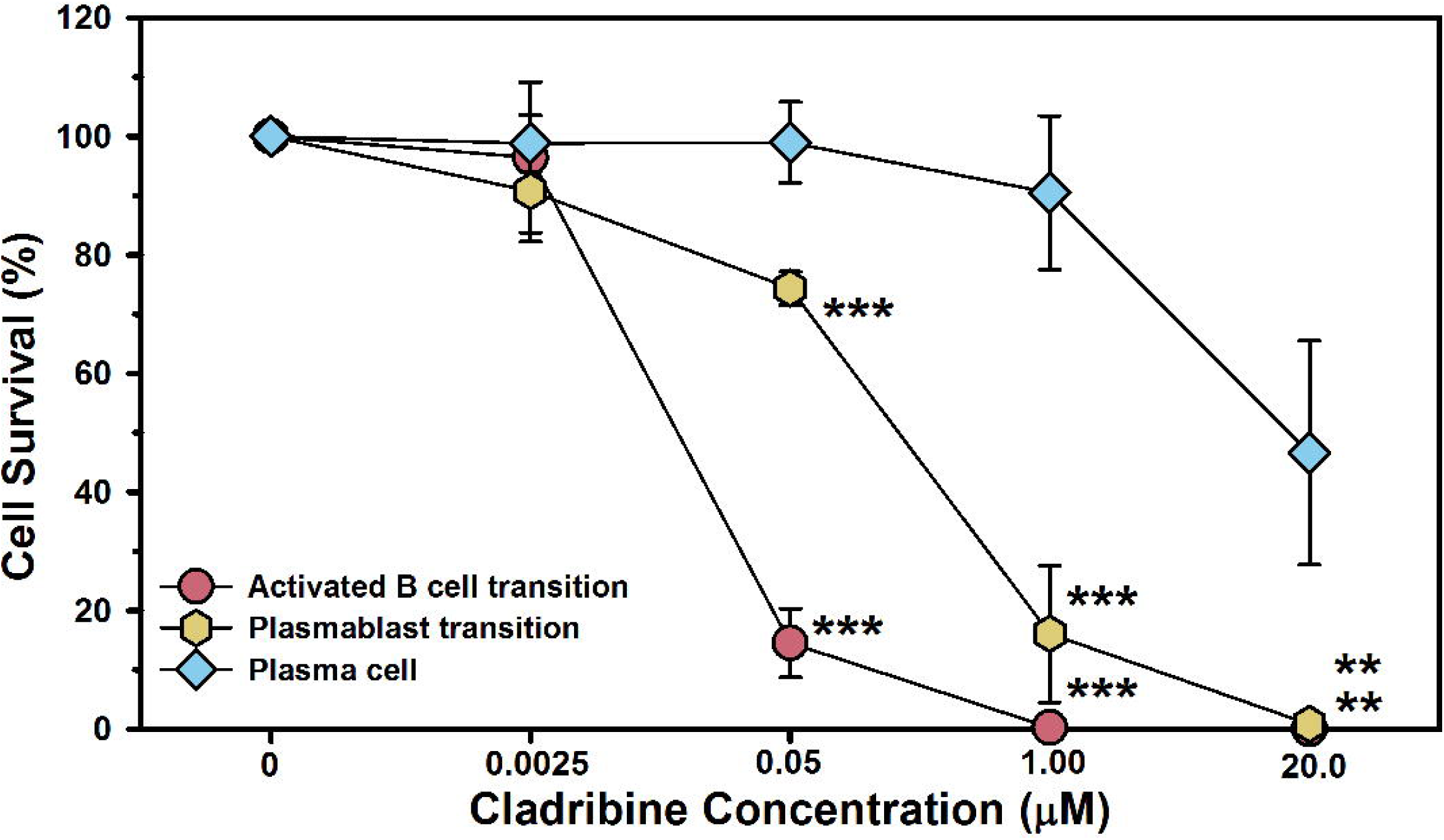
In vitro differentiated plasma cells are relatively resistant to the action of cladribine. Human peripheral blood total B cells were differentiated into plasma cells using cytokines, growth factors, B cell receptor binding and co-stimulation. The results represent the mean ± standard deviation (n=3) survival of cells following incubation of various concentrations of cladribine during the activated B cell to plasmablast transition [day 3-6], the plasmablast to plasma cell transition [day 6-8] and following plasma cell development [day 13-15]. ** P<0.01 ***P≤0.001 compared to survival of plasma cells [One-way Analysis of Variance].

## DISCUSSION

Given the ability of cladribine to penetrate the CNS, we originally hypothesised that cladribine would target intrathecal B cells and long-lived plasma cells to influence cOCB status and, thereby, potentially, disease progression, which is why we undertook CLADRIPLAS. However, as our understanding of the mode of action and direct cytotoxic effects of cladribine evolved ^11^, both microarray expression and *in vitro* cytotoxicity data contradicted our original hypothesis. However, consistent with the biology we uncovered, the number of people who were cOCB positive did not significantly drop following a first course of cladribine. Whilst our limited sample size precludes too deep an interpretation, the finding that cOCB disappeared in some but persist in most pwMS is consistent with previous observations using parenteral cladribine ^14^. Furthermore, our data supports the view that cOCB are not rapidly depleted by cladribine and is further enhanced by our recent data examining oral cladribine ^30^. We hypothesise this is due to limited plasma cell targeting by cladribine. Nonetheless, the biology is impacted such that whilst cOCB persisted in all nine participants 12-24 months after oral cladribine treatment, the number of OCBs was reduced in some people, alongside reduced free immunoglobulin kappa light chain levels ^30,31^. As seen with the differential sensitivities of B cell subsets to CD20-depletion in other conditions (Dziadkowiak et al. 2024), OCBs may represent a composite of antibodies produced by different antibody-secreting subsets. As such, they may be generated from long-lived plasma cells that may persist due to their resistance to cladribine and antibodies produced from short-lived plasmablasts that migrate to or are generated in the CNS following memory cell maturation, which are sensitive to the peripheral and central activity of cladribine, leading to loss of some bands as seen following oral cladribine ^30,31^).

Gene expression levels, notably of *DCK*, reflect the cellular depletion capacity of cladribine ^32^. Indeed it is evident that post-mitotic plasma cells are relatively insensitive to the effects of cladribine *in vitro,* which can be further augmented by an increase in ADA that can degrade cladribine to some extent ^32^. This sensitivity is consistent with the *in vivo* observations of persisting antibody and vaccination responses in cladribine-treated individuals (^33^; ^34^. This represents a significant safety element, indicating that childhood immunity to infections will persist. Indeed the steady state and peak CNS cladribine levels are likely to be several-fold lower than the concentration required to kill plasma cells completely.

Memory B cells express DCK and exhibit marked and long-lasting depletion ^35^; ^10^ consistent with the potent inhibition of MS as seen with cladribine and other active treatments of pwMS ^11^; ^32^. However, the level of B cell depletion in this study was less marked than anticipated and may relate to starting levels influenced by previous immunotherapies. Naïve B cells likewise exhibit DCK and are also sensitive to cladribine. However, in contrast to memory B cells they rapidly repopulate following depletion and drug elimination ^36^; ^10^. This facilitates responses to new infections thus contributing to the favourable safety profile of this agent ^37^. Persistence of established B cell immunity is also suggested from the observations that clonally expanded B cell repertoires persist despite treatment ^38^. In some individuals, disease returns as the immune system reconstitutes following depletion, but in most pwMS, the balance of immune regulation seems to favour effective long-term disease control ^32^ ^15^.

The reasons why OCB may be affected in some people more than others is not clear, but this may relate to the location of the individual plasma cell niches, local drug concentrations, cytokine levels supporting the plasma cell niches and degree of elimination of CNS inflammation. There is biological variation in the peripheral blood and, hence, intracerebral levels of cladribine, and the degree of lymphocyte depletion among individuals ^29,39^. There may also be differing levels of CNS penetration of cladribine due to varied expression and activity of endothelial exclusion transporters, such as the adenosine triphosphate binding cassette G2 (Breast cell cancer resistance protein) that controls the influx of cladribine and other molecules into the brain ^29,40^. However, incomplete removal of cOCB with time has also been noted following haematopoietic stem cell therapy (HSCT) and treatment with other peripheral immune inhibitions ^41–44^. It may, therefore, be more likely that cOCB levels are influenced by the removal of circulating, CNS-penetrating B cell precursors for antibody-secreting cells ^43,45^ or limiting the presence of cells providing cell contact and growth and survival factors required to maintain plasma cell niches. Simple inhibition of CNS B cells, possibly secondary to relapsing disease control, may not be sufficient to cause loss of cOCB ^45,46^.

Cladribine affects peripheral and CNS plasmablasts, and thus short-lived antibody-producing cells, as found in the blood ^10,47,48^. Direct activity on long-lived plasma cells, which are rarely found in the peripheral blood ^47^, is considered less likely given the notable downregulation of *DCK* and cladribine insensitivity as shown here. As such, circulating plasmablasts (CD20^-^, CD27^+^, CD38^+^) lose *DCK* during maturation and circulating plasmablast/plasma cells are only partially deleted (Median 66.6% depletion at 3 months (IQR 33.7-82.3 after oral cladribine. n=200. NCT03364036) compared to the naïve (Median 75.9% (IQR 66.2-84.2%) depletion) and memory B cell (Median 92.7% (IQR 86.80-94.1) depletion) pools ^10,47,48^. However, when examined following oral cladribine, we and others have shown that CD138^+^ plasma cells, which exhibit more limited DCK and cladribine sensitivity than plasmablasts, indeed also show more limited depletion than (CD10-, CD19+, CD38+) plasmablasts and memory B cells ^47^; ^30^. This is a valuable trait in terms of safety. As such, this view is consistent with the observations of limited hypogammaglobulinemia, maintenance of circulating immunoglobulin levels and *i n v i* v*v*ac*o*cination responses following cladribine treatment ^10,33,34,49^. Whilst our original hypothesis required modification, this study further highlights consistent biology that explains the efficacy and safety of cladribine in MS. We demonstrate a better understanding of the mechanisms of action of cladribine, a highly effective agent for pwMS. This mechanism largely equates to tissue expression of DCK and provides a robust biomarker for cladribine activity *in vivo*. We are confident this data will be replicated and expanded upon.

### Limitations

We used SClad, rather than the oral formulation (Mavenclad®) licensed for pwMS. However, we and others before us have shown that the two formulations are pharmacodynamically very similar ^13,29,32^. The dose of the licensed oral formulation was modelled on parenteral cladribine data ^29^. Likewise, the dosing schedule of CLADRPLAS was based on the pivotal CLARITY ^50^ study, including allowance for the enhanced bioavailability of subcutaneous cladribine ^13,29^. We therefore expect that the information shown in this study, which shows that the influences on plasma cells are much more limited than on memory B cells, applies to the licensed formulation. Indeed, consistent biology is emerging in relation to the effects on therapy ^29,50^; immunodepletion of cell subsets ^10,35^ and also vaccination and OCB responses ^30^. The small number of people recruited to the treatment arm in CLADRIPLAS was in part due to the COVID-19 pandemic. However, our conclusions are supported by the recently completed CLAD-B study ^30^.

## Data Availability

Data and supplementary materials are available on request.

## Acknowledgements.

The authors would like to thank the Multiple Sclerosis Society of Great Britain & Northern Ireland and Cancer Research UK for their support. Diagrams created with Biorender.com.

