## Supplementary figures and images for "Effects of cladribine on intrathecal and peripheral B and plasma cells"

### Supplementary Figure 1

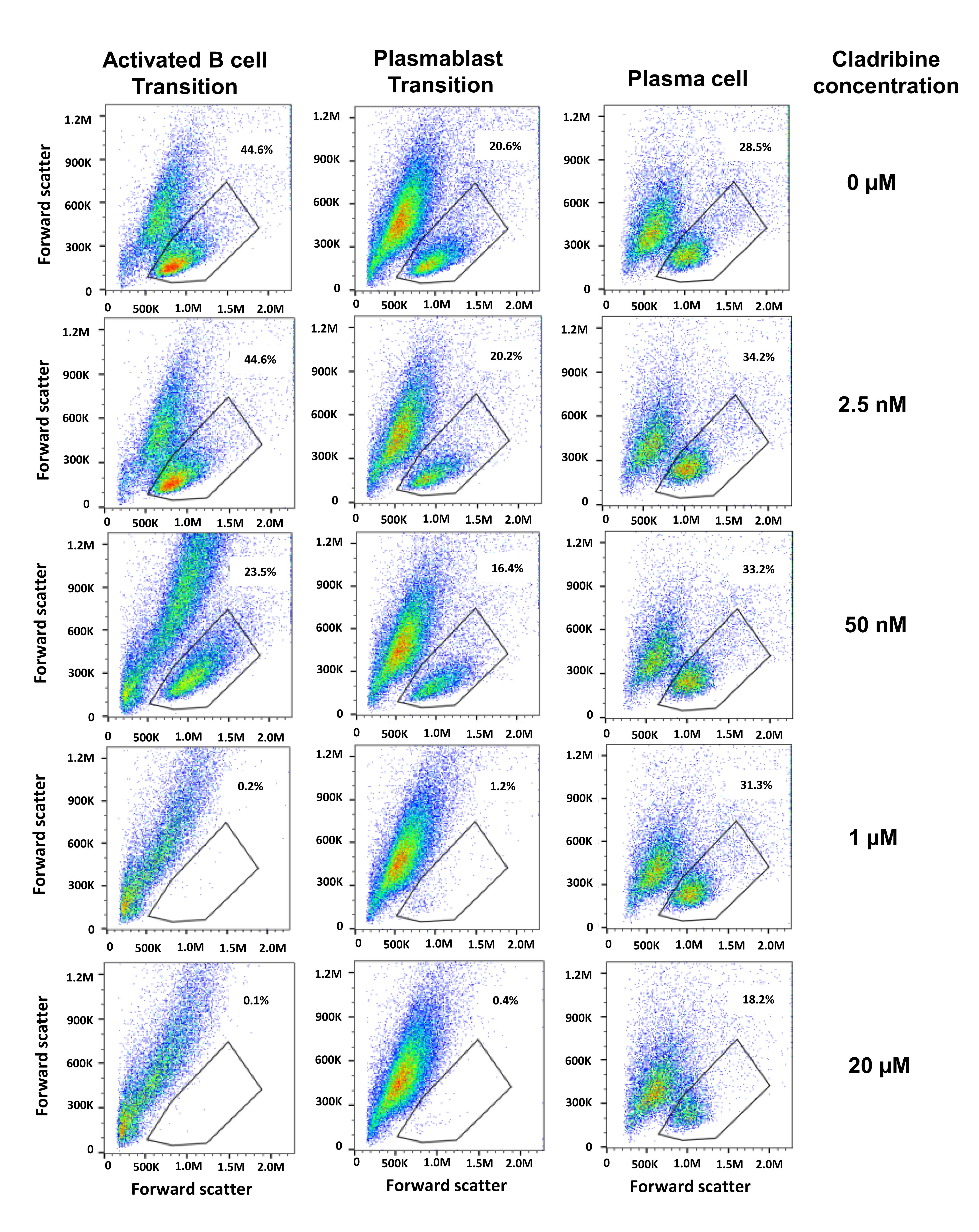

### Supplementary Figure 2

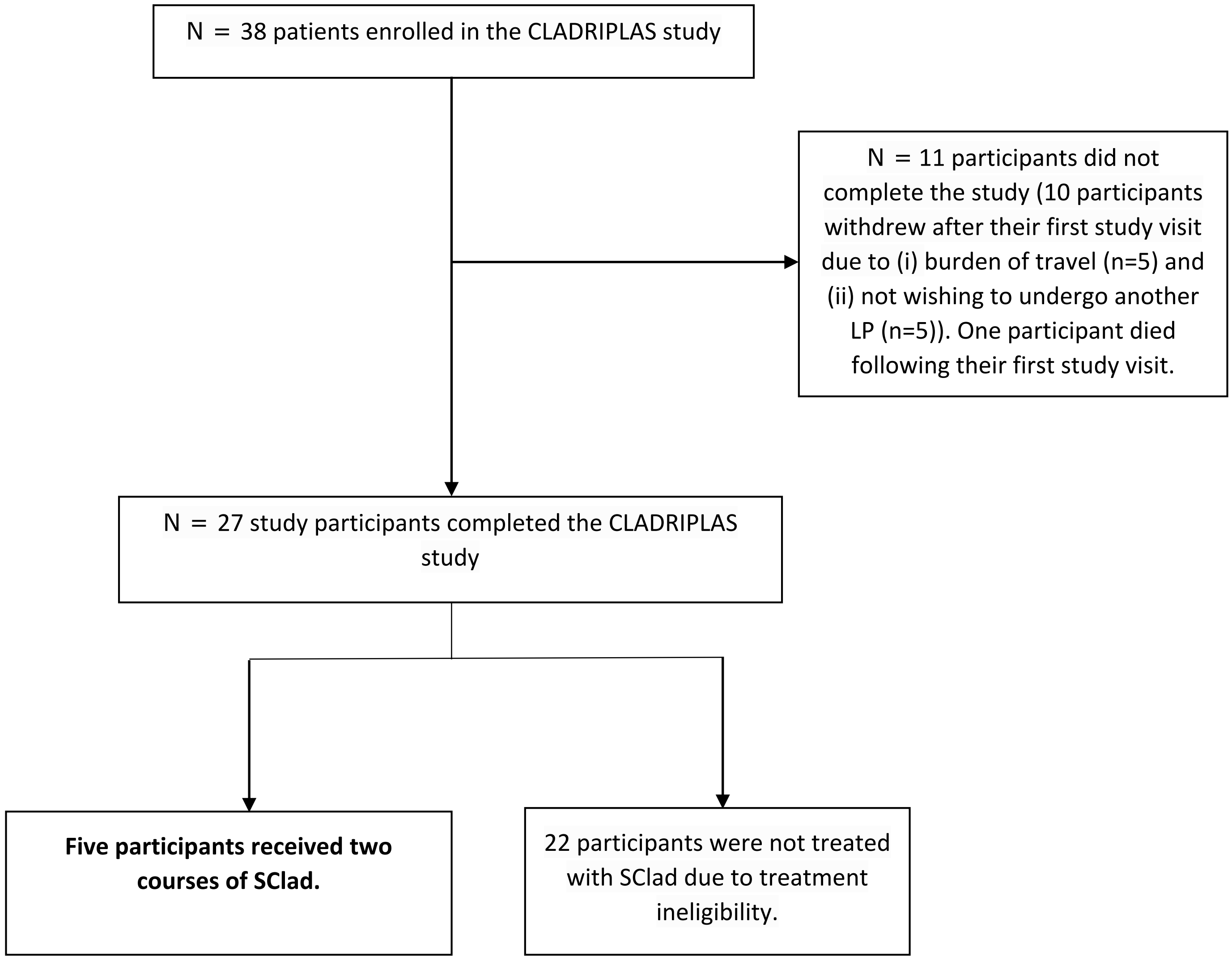
